# Histopathologist Features Predictive of Diagnostic Concordance at Expert Level Amongst a Large International Sample of Pathologists Diagnosing Barrett’s Dysplasia Using Digital Pathology

**DOI:** 10.1101/19000174

**Authors:** Myrtle J. van der Wel, Helen G. Coleman, Jacques JGHM Bergman, Marnix Jansen, Sybren L. Meijer, on behalf of the BOLERO working group

## Abstract

**Objective:** Guidelines recommend expert pathology review of Barrett’s oesophagus (BO) biopsies that reveal dysplasia, but there are no evidence-based standards to corroborate expert reviewer status. We investigated BO concordance rates and pathologist features predictive of diagnostic discordance amongst a large international cohort of gastrointestinal pathologists to develop a quantitative model of BO expert review.

**Design:** Pathologists (n=55) from over 20 countries assessed 55 digitised BO biopsies from across the diagnostic spectrum, before and after viewing matched p53 immunohistochemistry. Extensive demographic and clinical experience data were obtained via online questionnaire. We calculated discordance rates and applied multivariate regression analyses to identify predictors of concordance.

**Results:** We recorded over 6,000 individual case diagnoses. Of 2,805 H&E diagnoses, we found excellent concordance (>70%) for non-dysplastic Barrett’s oesophagus (NDBO) and high-grade dysplasia (HGD), and intermediate concordance for low-grade dysplasia (LGD, 42%) and indefinite for dysplasia (IND, 23%). Major diagnostic errors (i.e. NDBO overinterpreted as LGD/HGD or vice versa) were found in 248 diagnoses (8.8%), which reduced to 8.3% after viewing p53 labelled slides. At least 5 years of professional experience was protective against major diagnostic error for H&E slide review (OR 0.48, 95%CI 0.31-0.74). Working in a district general hospital was associated with increased odds of major diagnostic error (OR 1.76, 95%CI 1.15-2.69), however this was neutralised when pathologists viewed p53 labelled slides, suggesting a beneficial impact of p53 immunohistochemistry for this group.

**Conclusion:** We have developed an evidence-based quantitative model of BO histopathology diagnosis at expert consensus level that will inform guideline development.

## INTRODUCTION

Barrett’s oesophagus (BO) is a premalignant condition, which predisposes to esophageal adenocarcinoma (OAC), with a reported annual conversion rate of 0.1 - 0.2%. ^1-3^ BO is defined histopathologically as the replacement of normal stratified squamous epithelial lining of the distal oesophagus with columnar epithelium that can contain intestinal metaplasia. The implementation of formal surveillance strategies and widespread adoption of endoscopic treatment techniques, such as endoscopic resection and ablation for dysplastic BO, have led to a surge in diagnostic pathology workload. The goal of endoscopic surveillance and biopsy verification is objective risk stratification for patients according to their perceived progression risk to OAC.

Previous studies have revealed, however, that diagnostic reproducibility (inter-observer agreement) amongst pathologists grading dysplastic BO biopsy material is moderate to poor, even amongst expert reviewers (**Supplementary Table 1**). ^4-17^ Previous work from our group has shown that central pathology review by a dedicated panel within the context of prospective intervention trials failed to confirm an initial diagnosis of low-grade dysplasia (LGD) in over three-quarters of cases submitted for panel review. On follow up, cases that had been downgraded to non-dysplastic BO (NDBO) revealed a nominal progression risk of about 0.5% per patient/year, whilst cases that had been confirmed LGD on central review showed a progression risk of about 10% per patient/year. These data clearly attest to the clinical return of dedicated pathology review. ^18 19^ International BO management guidelines now mandate histopathology review of all BO biopsy cases found to reveal dysplasia by an independent expert pathologist. ^20 21^ However, whilst major society guidelines have qualitatively defined an expert BO pathologist as ‘a pathologist with a special interest in BO-related neoplasia who is recognised as an expert in this field by their peers’, we lack firm evidence-based standards to corroborate expert reviewer status. ^21-26^ This now represents an acute unmet need as these considerations also carry important medico-legal implications.

Recently, the US Food and Drug Administration has approved the use of whole slide imaging (WSI) for primary diagnostic use. ^27^ The advantages of WSI are numerous and include simultaneous assessment by multiple pathologists, streamlined expert consultation, and digital image analysis. It is expected that digital pathology will rapidly gain widespread acceptance in the coming years, in particular in the context of distant case review. A number of large-scale diagnostic consensus studies have been performed, which have broadly suggested that the diagnostic discordance rate between pathologists using digital slide review is non-inferior to conventional glass slide diagnosis. ^28-30^ However, these studies generally examined a large number of diagnostic categories without focusing on a particular category of known diagnostic discordance such as Barrett’s dysplasia. Establishing the validity of this new technology to BO histopathologic workup is therefore a clear priority.

Here we set out to develop quantitative standards of expert reviewer status for guideline development purposes using massive online digital pathology reporting. We define expert reviewer status as evidence of diagnostic concordance on a par with consensus within an expert review panel, acknowledging that, in lieu of an objective biomarker of progression risk, there will be diagnostic variation amongst expert pathologists. We collected extensive demographic information of participating pathologists to understand operator-dependent predictors of diagnostic variation.

## METHODS

### Ethical considerations

This study utilised anonymised archived formalin-fixed, paraffin embedded material and did not require approval from the relevant Institutional Ethics Committee under applicable local regulatory law (‘Code of conduct’, FEDERA).

### Assessors

Sixty-five gastrointestinal pathologists worldwide were approached to join this study through either professional gastrointestinal pathology working groups or direct professional contacts. Fifty-nine pathologists responded positively to our enquiries and were recruited to this study of which 51 pathologists completed the entire case set of 55 H&E-stained and 55 matching p53 immunohistochemistry (IHC) labelled slides (110 slides total). These 51 pathologists are henceforth referred to as participating pathologists. Participating pathologists received emails detailing the study objectives and were provided with personal log-in credentials to the purpose-built online scoring environment described below. Lead study author (MvdW) provided assistance with participating pathologists’ log-in queries, evaluated study progress, and chaired the panel consensus meeting.

Four BO pathologists (including two study authors, MJ and SM) with extensive experience in BO dysplasia assessment reviewed all slides as a reference pathologist panel. This group has successfully collaborated on previous BO intervention studies where patient outcome has been evaluated prospectively ^18 19 31-37^ as well as on the Amsterdam Barrett’s Advisory Committee. ^31^ These pathologists are henceforth referred to as reference pathologists.

### Slide selection and scanning

The lead study author selected a representative case-mix of 55 BO biopsy cases from across the diagnostic spectrum (**Supplementary Table 2**). Inclusion criteria were: diagnosis confirmed by a second gastrointestinal pathologist; documented clinical follow-up of at least one year available; and tissue block available. Per case, immunohistochemical staining for p53 was performed using a Ventana Benchmark XT autostainer (Ventana Medical Systems, Tucson, AZ). Antigen retrieval was performed with CC1 mild. P53 was detected with p53 Antibody (Mouse DO-7 + BP 53-12, Thermo Scientific) and the sections were incubated in a 1:500 dilution for 32 min at room temperature. Bound antibody was detected using the Biotin free Ultraview Universal DAB Detection Kit (Roche Diagnostics) and slides were counterstained with Hematoxylin (Roche Diagnostics). ^38^ One H&E slide and one consecutive section p53 IHC slide were digitised from each case using a scanner with a 20x microscope objective (Slide, Olympus, Tokyo, Japan). Scans were checked for focus and acuity by the study coordinator and re-scanned if necessary. Subsequently, slides were anonymised, randomised, renamed, and stored on a secure server. The ‘Digital Slidebox 4.5’ (https://dsb.amc.nl/dsb/login.php, Slidepath, Leica Microsystems, Dublin, Ireland) virtual slide viewing software was used to evaluate the digital slides during the study.

### Electronic scoring environment

Template electronic Case Record Forms (CRFs) were custom built within a web-based software tool designed to capture clinical study data (OpenClinica v3.6, an open source CTMM TraiT project, LLC, Waltham, USA). One CRF consists of an extensive questionnaire documenting pathologist characteristics such as age, sex, host institution, and experience in reporting BO biopsies and digital pathology (full questionnaire details in **Supplementary Table 3**). The second CRF was built to record individual case diagnoses. Importantly, this second CRF consists of separate parts to record H&E and H&E plus p53 IHC slide diagnoses independently. The first part of the case diagnosis CRF contains a dynamic URL link to the scanned H&E slide and includes questions about the slide quality and diagnosis, and whether the assessor feels they require a p53 IHC slide. Importantly, the second part of the templated CRF that contains a dynamic link to the p53 IHC slide alongside the matching H&E slide, only opens after the study pathologist has completed assessment of the H&E-stained slide and saved their case diagnosis for this slide. This second part of the templated CRF, in addition to a dynamic link to the matching p53 IHC slide, again included corresponding slide assessment questions.

### Digital case assessments

Reference and participating pathologists were asked to assess each case, according to the modified Vienna classification for gastrointestinal neoplasia. ^39 40^ Reference pathologists first assessed all cases individually and completed the questionnaire. An online consensus meeting was then convened after a two-month wash out period to discuss discrepancies and produce reference diagnoses for each of the 110 assessments (55 H&E-stained slides and 55 matching p53 IHC). The panel assessment was taken forward as the reference diagnosis without further discussion if reference panel members achieved a majority diagnosis (i.e. concordance between either 3 out of 4 or 4 out of 4 pathologists) on a case directly from their independent scoring. Group discussions were held between these four pathologists to review and discuss cases for which there was no majority diagnosis to mimic real-world practice. The discrepancies where a majority diagnosis had not been reached after individual slide review encompassed 21 cases based on H&E slide viewing, and 13 cases based on the p53 IHC slide. These cases were reviewed during the panel discussion (21 H&E slides reviewed without matching p53 IHC slide, and 13 cases with H&E-stained slide and matching p53 IHC) to arrive at a consensus diagnosis for all 110 assessments.

From the case assessments by the participating pathologists two p53 IHC case assessments were inadvertently left blank by individual participating pathologists (one each) after evaluating the case H&E slide. Results from the matching H&E slides were imputed as p53 case diagnosis in these cases, based on the H&E slide score, corresponding to 2 HGD diagnoses.

### Statistical analysis

Characteristics of the four reference pathologists and the 51 participating pathologists were compared informally. We examined the overall concordance of the study pathologists compared to the consensus reference diagnosis per case. This process was conducted for each of the four individual members of the reference panel against the final consensus diagnosis of this panel, as well as for the overall sample of 51 pathologists against the consensus diagnosis. Per pathologist scores were not calculated, since we aimed to study the cohort behavior rather than the individual pathologist. Concordance was initially compared based on four relevant diagnostic categories (NDBO, IND, LGD, HGD), and then compared based on three relevant diagnostic categories (NDBO, IND, LGD or HGD) to reflect the fact that HGD and LGD are now treated endoscopically in some settings. ^32^ We calculated 95% CIs for overall concordance and per diagnostic category. Since this cohort was strongly enriched for dysplasia, we did not use kappa statistics, since these are less reliable when cross tables are skewed.

To evaluate the severity of discordant interpretations across the cohort of participating pathologists, we then reclassified all discordant assessments as either major or minor discordances. Major overinterpretation is defined as NDBO reference diagnosis overinterpreted as either LGD or HGD, whereas, vice versa, major underinterpretation is LGD or HGD reference diagnosis underinterpreted as NDBO by the participating pathologist. These discordant interpretations would bear major consequences in clinical practice. All other discordant interpretations were classified as minor discordant interpretations. A tabular overview of interpretation classifications as major or minor is shown in **Supplementary Table 4**. Since both major overinterpretation and major underinterpretation can have negative implications for patient management, these were further combined for the purposes of some analyses, as indicated.

Unadjusted logistic regression analyses were then conducted to identify any pathologist characteristics that were associated with overall and major over or underinterpretation of BO cases, compared to the consensus diagnosis. Considering that age and professional experience are inextricably linked, we evaluated individual combinations of age and experience for odds of major over and underinterpretations, and combined these into three categories in whom similar odds ratios were observed (**Supplementary Table 5**). Forward selection of significant factors was used to create multivariable-adjusted logistic regression models of characteristics associated with misinterpretation. Although routine use of p53 immunohistochemistry was not associated with diagnostic errors, this was retained in multivariate models for p53 stained slides. All statistical analyses were performed using Stata version 14.2 (StataCorp., College Station, TX, USA).

## RESULTS

### Study design

This study is based on assessments of digitised slides to investigate diagnostic concordance of BO biopsies amongst a large and heterogeneous sample of gastrointestinal pathologists. We investigated rates and features predictive of diagnostic concordance amongst these pathologists, with a particular focus on the demographic characteristics of the pathologists, the impact of viewing p53 labelled slides alongside H&E-stained slides, and on features associated with major diagnostic discordance that would negatively impact upon patient stratification and treatment pathways. The purpose of this study was to build a quantitative model of expert BO pathologist review characteristics, and to provide practical recommendations that could minimize errors in the interpretation of BO biopsies in the routine setting.

The study flowchart is shown in **Figure 1A**. All pathologists first filled out a baseline questionnaire for detailed demographic and clinical experience data. Pathologists then assessed the 110 digitised slides (55 H&E slides and matching p53 IHC) and recorded their answers on dedicated electronic CRFs. As detailed in the methods section, diagnostic entries were recorded after viewing the H&E-stained slide and again after the matched p53 IHC was revealed alongside the case H&E slide.

**Figure 1:**
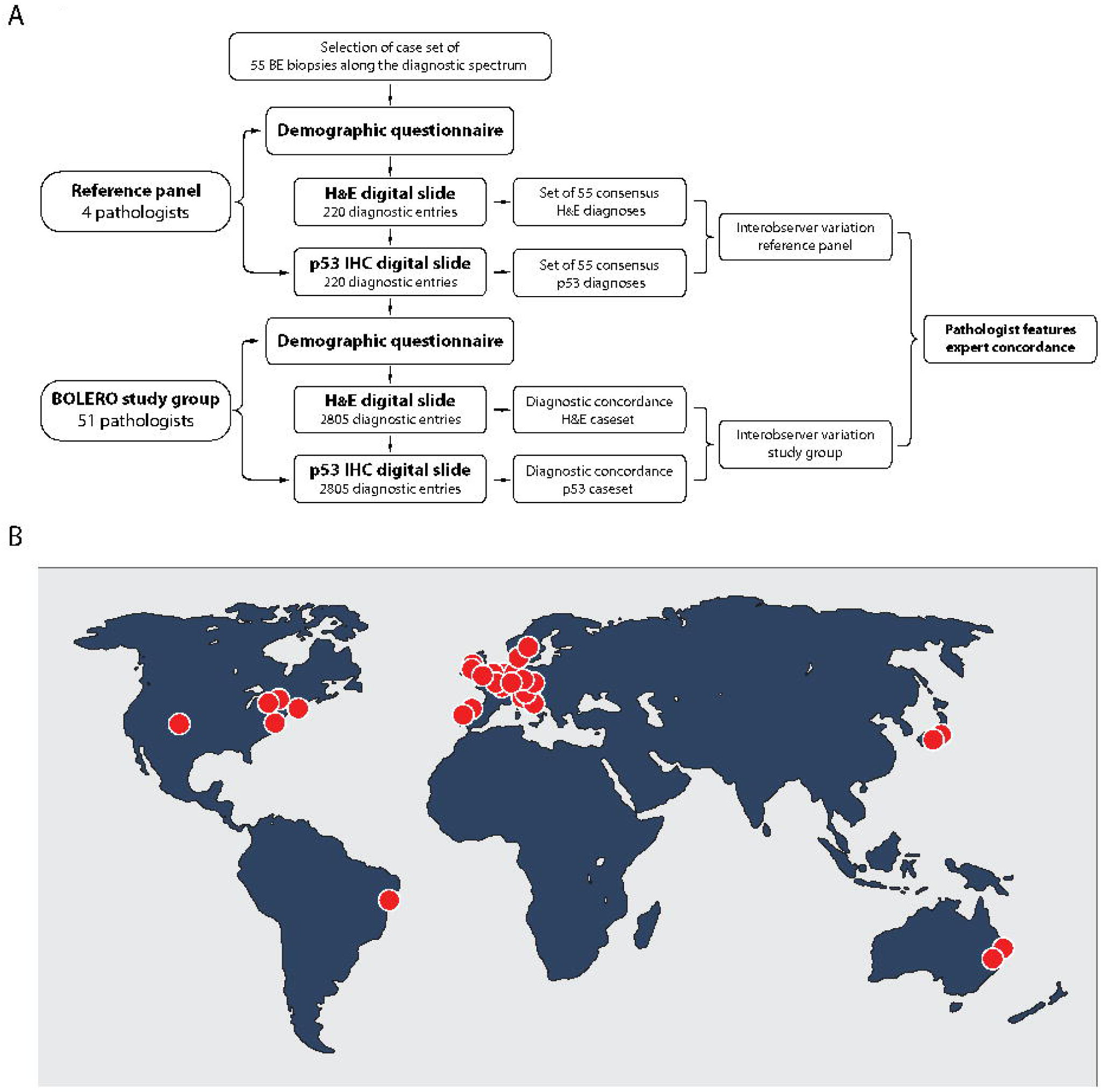
Study design and study participants. A) Fifty-five representative BO biopsies with H&E slide and consecutive p53 IHC were collected and scanned for digital diagnostic review. Each pathologist on the study first completed a detailed demographic questionnaire (Supplementary Table 3). Pathologists then assessed 55 biopsy cases whereby diagnostic entries on H&E slide alone and after revealing matched p53 IHC were recorded separately allowing detailed insight into the added benefit of p53 IHC on diagnostic agreement. Reference diagnoses were established after consensus panel meeting. Within-group interobserver agreement was established for reference panel (n=4) and participating pathologists (N=51) and multivariate regression analyses were carried out to interrogate demographic predictors of diagnostic concordance, as detailed in the text. B) Map showing geographical dispersion of pathologists participating in the BOLERO study.

The entire study set was completed by fifty-five pathologists working in over 20 countries and 5 continents (**Figure 1B**). Of these fifty-five pathologists, 4 pathologists with extensive and published experience in BO dysplasia assessment were designated beforehand as reference pathologists. ^18 19 32 41 42^ In sum, with 55 pathologists reviewing 55 biopsy cases, each of which includes one H&E-stained slide and a matched p53 IHC, this generated a massive dataset of over 6,000 case diagnoses as input data for our Barrett’s digital pathology (BOLERO) consensus study, one of the largest digital pathology consensus studies reported thus far. Case diagnoses were compared to reference diagnoses and we searched for pathologist demographic features that predict diagnostic consensus at expert level.

### Patient characteristics of BO biopsy samples

Patient characteristics of the sample biopsies are shown in **Supplementary Table 2**. Of these patients, 94.5% was male (52/55). The median age at diagnosis was 65, the median BMI was 27, the median BO segment length was Circumferential (C) 4 cm, Maximum (M) 5 cm. Patients had a history of smoking in 63.6% of cases (35/55), a history of heartburn symptoms in 89% of cases (49/55), and used anti-reflux medication in 96.4% of cases (53/55).

### Pathologist characteristics

Baseline characteristics of the pathologists taking part in the study are displayed in **Table 1** and **Supplementary table 6**. Participating pathologists represented a heterogeneous sample comprising a wide range of ages, workplace settings (academic teaching, private and/or district general hospital settings) and years of professional experience. Just over 50% of participating pathologists reported dedicated fellowship experience, whilst the majority (72%) worked in a large laboratory with ≥10 pathologist colleagues. The most commonly reported guidelines to which pathologists adhered were North American, British, or Japanese, however a quarter of pathologists reported using other guidelines in their clinical practice. Two thirds of participating pathologists self-identified as expert gastrointestinal pathologists. Note that although pathologists were approached through international working groups, no effort was made to purposely recruit experts onto the study. Pathologists also reported on other parameters and working practices in their laboratories, such as typical numbers of BO cases reported per week, confidence and enjoyment in reporting BO, reporting of endoscopic resection specimens, frequency of adjunct p53 IHC use in BO reporting, participation in double-reporting, multi-disciplinary team meetings, and use of WSI, as well as typical interactions and perceptions of practices of their endoscopy colleagues (**Table 1 and Supplementary table 6**). Participating and reference pathologists were generally well matched for age ranges and professional experience although all four reference pathologists were male, whereas 22 of 51 (43.1%) participating pathologists in the larger cohort were female.

**Table 1:**
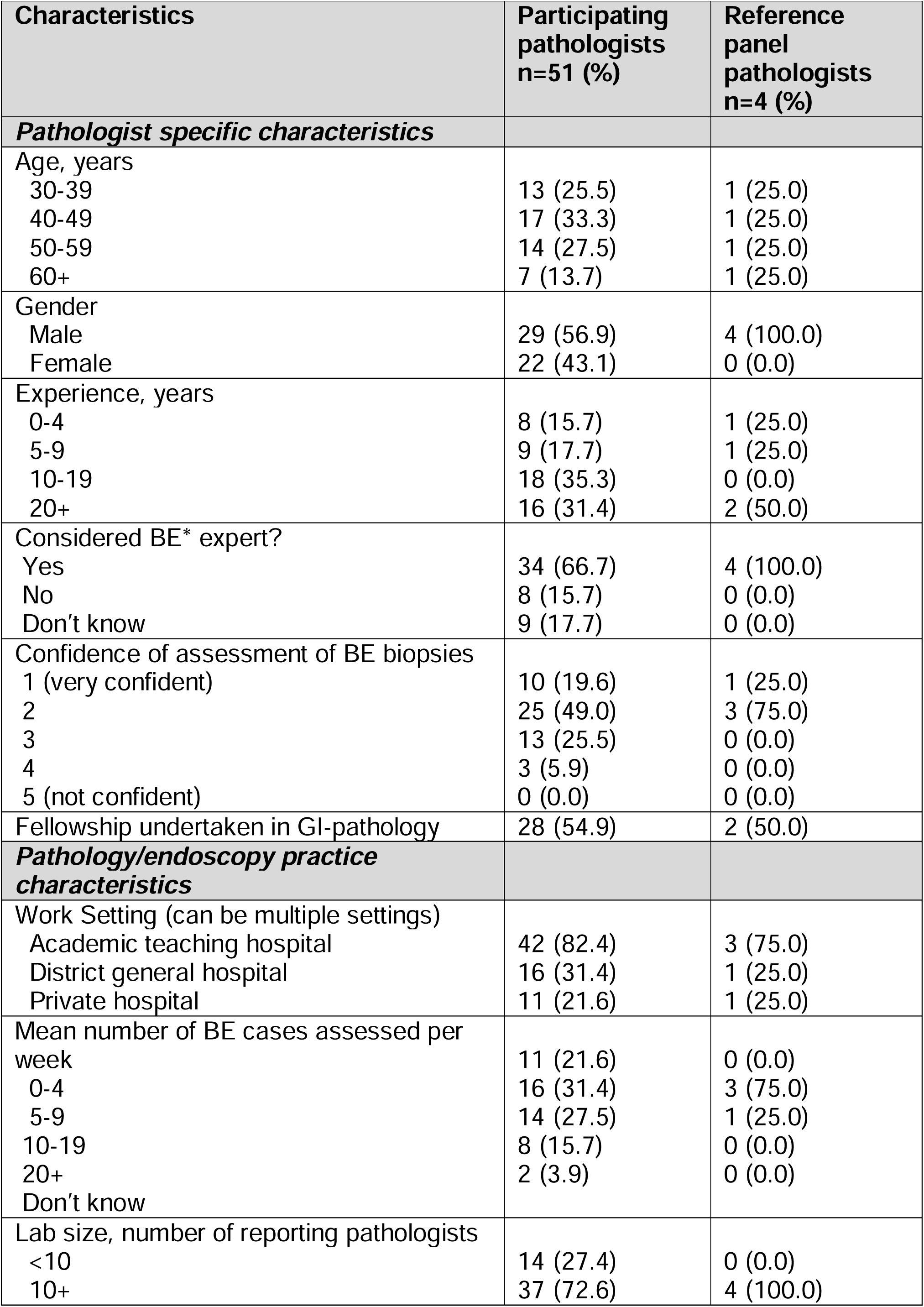
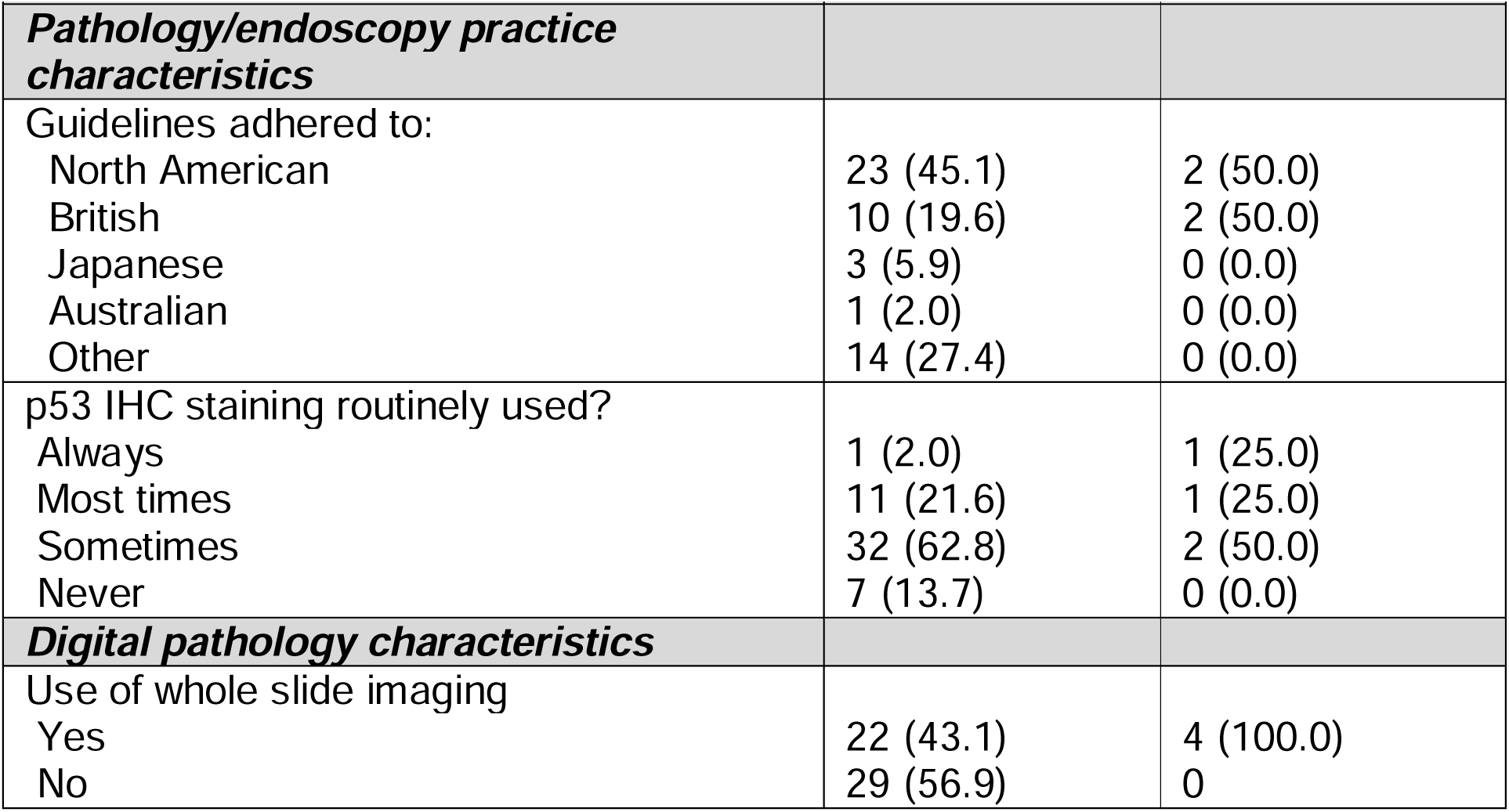
Demographics of pathologists reporting in the BOLERO study.

| **Characteristics** | **Participating pathologists**<br>**n=51 (%)** | **Reference panel pathologists**<br>**n=4 (%)** |
| --- | --- | --- |
| ***Pathologist specific characteristics*** |  |  |
| Age, years |  |  |
| 30-39 | 13 (25.5) | 1 (25.0) |
| 40-49 | 17 (33.3) | 1 (25.0) |
| 50-59 | 14 (27.5) | 1 (25.0) |
| 60+ | 7 (13.7) | 1 (25.0) |
| Gender |  |  |
| Male | 29 (56.9) | 4 (100.0) |
| Female | 22 (43.1) | 0 (0.0) |
| Experience, years |  |  |
| 0-4 | 8 (15.7) | 1 (25.0) |
| 5-9 | 9 (17.7) | 1 (25.0) |
| 10-19 | 18 (35.3) | 0 (0.0) |
| 20+ | 16 (31.4) | 2 (50.0) |
| Considered BE* expert? |  |  |
| Yes | 34 (66.7) | 4 (100.0) |
| No | 8 (15.7) | 0 (0.0) |
| Don't know | 9 (17.7) | 0 (0.0) |
| Confidence of assessment of BE biopsies |  |  |
| 1 (very confident) | 10 (19.6) | 1 (25.0) |
| 2 | 25 (49.0) | 3 (75.0) |
| 3 | 13 (25.5) | 0 (0.0) |
| 4 | 3 (5.9) | 0 (0.0) |
| 5 (not confident) | 0 (0.0) | 0 (0.0) |
| Fellowship undertaken in GI-pathology | 28 (54.9) | 2 (50.0) |
| ***Pathology/endoscopy practice characteristics*** |  |  |
| Work Setting (can be multiple settings) |  |  |
| Academic teaching hospital | 42 (82.4) | 3 (75.0) |
| District general hospital | 16 (31.4) | 1 (25.0) |
| Private hospital | 11 (21.6) | 1 (25.0) |
| Mean number of BE cases assessed per week |  |  |
| 0-4 | 11 (21.6) | 0 (0.0) |
| 5-9 | 16 (31.4) | 3 (75.0) |
| 10-19 | 14 (27.5) | 1 (25.0) |
| 20+ | 8 (15.7) | 0 (0.0) |
| Don't know | 2 (3.9) | 0 (0.0) |
| Lab size, number of reporting pathologists |  |  |
| <10 | 14 (27.4) | 0 (0.0) |
| 10+ | 37 (72.6) | 4 (100.0) |
| ***Pathology/endoscopy practice characteristics*** |  |  |
| Guidelines adhered to: |  |  |
| North American | 23 (45.1) | 2 (50.0) |
| British | 10 (19.6) | 2 (50.0) |
| Japanese | 3 (5.9) | 0 (0.0) |
| Australian | 1 (2.0) | 0 (0.0) |
| Other | 14 (27.4) | 0 (0.0) |
| p53 IHC staining routinely used? |  |  |
| Always | 1 (2.0) | 1 (25.0) |
| Most times | 11 (21.6) | 1 (25.0) |
| Sometimes | 32 (62.8) | 2 (50.0) |
| Never | 7 (13.7) | 0 (0.0) |
| ***Digital pathology characteristics*** |  |  |
| Use of whole slide imaging |  |  |
| Yes | 22 (43.1) | 4 (100.0) |
| No | 29 (56.9) | 0 |

### Case assessment overview

A total of 3,025 diagnoses were generated based on H&E-stained slide case review and another 3,025 diagnoses were recorded after viewing the matching p53 IHC slides for study cases (**Figure 2A and B**). The corresponding waterfall plots showing the ranked distribution of assessments reveal a gradual transition from NDBO examples with high interobserver concordance to HGD cases with similarly high interobserver concordance and diagnostic categories where concordance gradually transitions between these extremes. These plots also confirm that our case set includes representative biopsies from across the diagnostic spectrum of BO pathology. Relevant examples of study cases are shown in **Figure 2C**.

**Figure 2:**
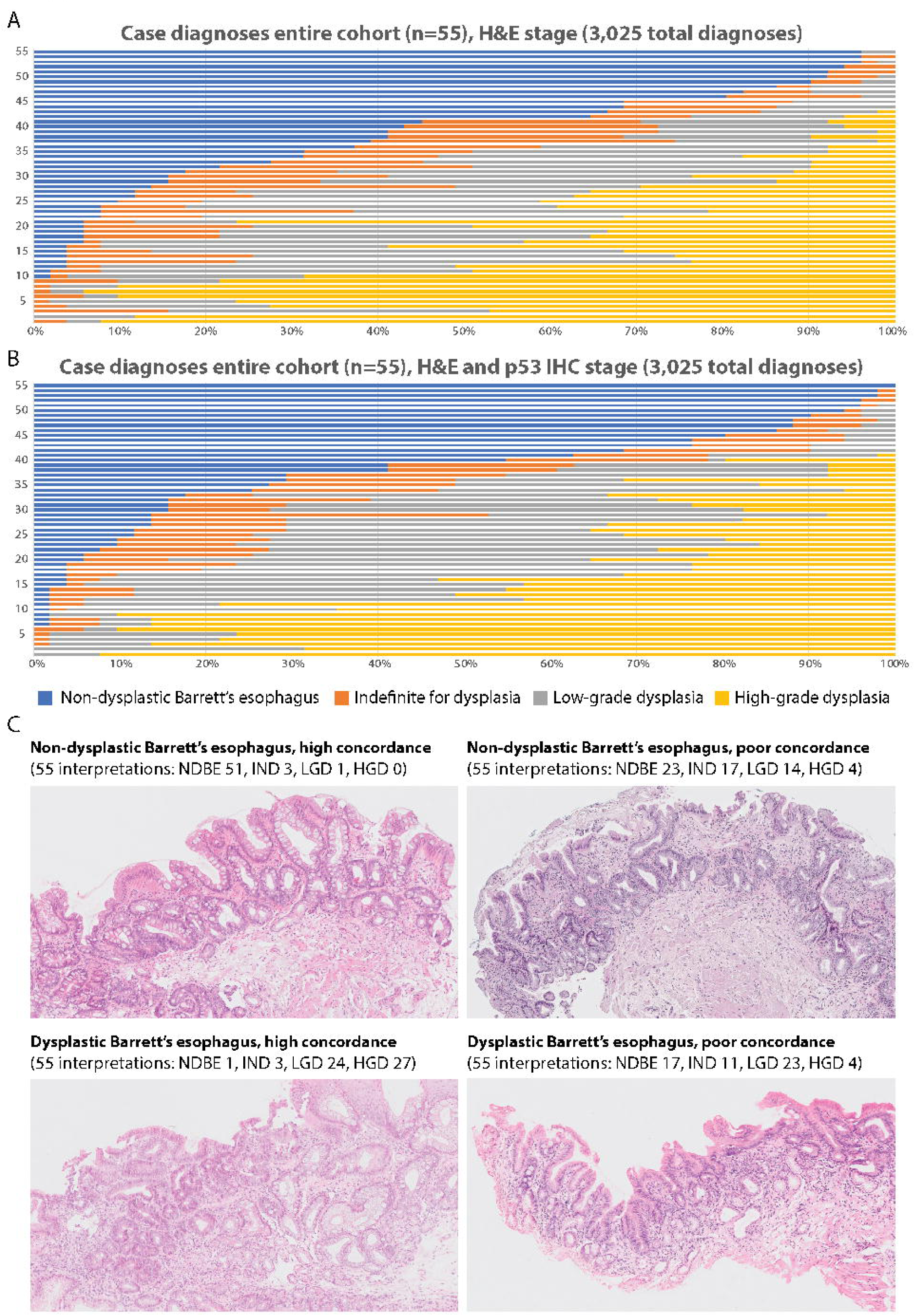
Diagnostic variation across the study cohort. A) Waterfall plot showing the ranked distribution of case assessments (n=3,025) based on H&E slides alone for the entire cohort of pathologists. X-axis shows diagnostic concordance in percentages and y-axis shows ranked cases 1-55. Color coding as in B. B) Same visualisation for case assessments (n=3,025) after revealing matched p53 IHC slide. C) Four representative examples of the study set. Consensus diagnosis and cohort diagnoses are shown.

### Concordance of reference pathologists vs. consensus diagnosis on H&E and p53 labelled slides

Consensus diagnoses were generated following panel review. The reference panel consensus diagnoses for the H&E-stained slide case review included 16 NDBO, 6 IND, 18 LGD, and 15 HGD case diagnoses. After the addition of matched p53 IHC and reference panel review a small number of cases were reclassified, including 1 NDBO diagnosis as LGD, 1 LGD diagnosis as NDBO, and 4 IND diagnoses as LGD, thus totaling 16 NDBO, 2 IND, 22 LGD and 15 HGD after p53 IHC slide review.

Individual consensus panel member diagnoses were then compared to the final consensus panel diagnosis to obtain concordance rates between the 4 reference pathologists. This revealed excellent diagnostic agreement when reporting NDBO, LGD and HGD on H&E-stained slides alone (84.4%, 65.3% and 78.3%, respectively), rising to 89.4% when LGD and HGD diagnoses were combined. After revealing the matching p53 IHC slide for the 55 cases, agreement further improved to 85.9% for ND, 72.7% for LGD, and 76.7% for HGD, rising to 91.9% when LGD and HGD were combined (**Supplementary Tables 7A and B**).

### Concordance of participating pathologists vs. consensus diagnosis on H&E and p53 stained slides

The complete set of 5,610 case assessments recorded by the 51 participating pathologists was then compared to the reference panel diagnoses to obtain concordance rates and compare diagnostic agreement within and between categories. The diagnostic agreement between 51 participating pathologists for H&E-stained slide diagnoses is depicted in **Figure 3A-C** and **Supplementary Figure 1A**, while concordance percentages are shown in **Table 2A**. We found excellent concordance between the participating pathologists for NDBO reference diagnosis cases (643 of 816 diagnoses; 78.8%) and HGD reference diagnosis cases (544 of 765 diagnoses; 71.1%). As expected, there was moderate concordance for LGD reference diagnosis cases (382 of 918; 41.6%) and poor concordance for IND reference diagnosis cases (70 of 306; 22.9%). However, if dysplastic assessments were grouped (i.e. combining LGD and HGD reference diagnosis cases) then 77.5% (1,305 of 1,683) of cases were concordant. Major over or underinterpretation was found in 8.8% of assessments (248 of 2,805 diagnoses).

**Table 2:** Cross table comparing the 51 participating pathologists’ diagnoses to the consensus derived reference diagnoses for 55 esophageal biopsy cases (a) on HE staining and (b) on HE and p53 IHC staining for 5,610 total case interpretations*. *Overall concordance for 1639/2805 diagnoses (58.4%, 95%CI 56.6-60.2%); increasing to 2018/2805 (71.9%, 95%CI 70.2-73.6%) when LGD and HGD were combined, **Note consensus reference panel results are scaled x51 to allow for comparison versus the 51 participating pathologists. Results represent 5,610 diagnoses in 55 oesophageal biopsy cases. ***Overall concordance for 1765/2805 diagnoses (62.9%, 95% CI61.1-64.7%); increasing to 2205/2805 (78.6%, 95%CI 77.1-80.1%) when LGD and HGD were combined.

|  | Consensus<br>reference<br>panel** | Participating pathologists'<br>individual diagnoses<br>(preconsensus) |  |  |  | % Concordance<br>(95% CI) |  |  |
| --- | --- | --- | --- | --- | --- | --- | --- | --- |
|  |  |  |  |  |  | Under-<br>interpretat<br>ion | Over-<br>interpretati<br>on | Concorda<br>nce |
| a. Before addition of p53 immunohistochemistry |  |  |  |  |  |  |  |  |
| Diagnosis |  | ND | IND | LGD | HGD |  |  |  |
| NDBO | 816 | 643 | 93 | 71 | 9 | / | 21.2<br>(18.4-24.0) | 78.8<br>(0.70-81.6) |
| IND | 306 | 59 | 70 | 110 | 67 | 19.2<br>(14.8-23.6) | 57.8<br>(52.3-63.3) | 22.9<br>(18.2-27.6) |
| LGD | 918 | 151 | 165 | 382 | 220 | 34.4<br>(31.3-37.5) | 24.0<br>(21.2-26.8) | 41.6<br>(38.4-44.8) |
| HGD | 765 | 17 | 45 | 159 | 544 | 28.9<br>(25.7-32.1) | / | 71.1<br>(25.6-32.2) |
| LGD or HGD | 1683 | 168 | 210 | 1305 |  | 22.5<br>(20.4-24.5) | / | 77.5<br>(75.5-79.5) |
| Total | 2805 |  |  |  |  |  |  |  |
|  | Consensus<br>reference<br>panel*** | Participating pathologists'<br>individual diagnoses<br>(preconsensus) |  |  |  | % Concordance<br>(95% CI) |  |  |
|  |  |  |  |  |  | Under-<br>interpretat<br>ion | Over-<br>interpretati<br>on | Concorda<br>nce |
| b. After addition of p53 immunohistochemistry |  |  |  |  |  |  |  |  |
| Diagnosis |  | ND | IND | LGD | HGD |  |  |  |
| NDBO | 816 | 684 | 74 | 53 | 5 | / | 16.2<br>(13.7-18.7) | 83.8<br>(81.3-86.3) |
| IND | 102 | 36 | 24 | 27 | 15 | 35.3<br>(26.0-44.6) | 41.2<br>(31.6-50.8) | 23.5<br>(15.3-31.7) |
| LGD | 1122 | 153 | 178 | 516 | 275 | 29.5<br>(26.8-32.2) | 24.5<br>(22.0-27.0) | 46.0<br>(43.7-49.5) |
| HGD | 765 | 21 | 38 | 165 | 541 | 29.3<br>(26.1-32.5) | / | 70.7<br>(67.8-73.9) |
| LGD or HGD | 1887 | 174 | 216 | 1497 |  | 20.7<br>(18.9-22.5) | / | 79.3<br>(77.5-81.1) |
| Total | 2805 |  |  |  |  |  |  |  |

**Figure 3:**
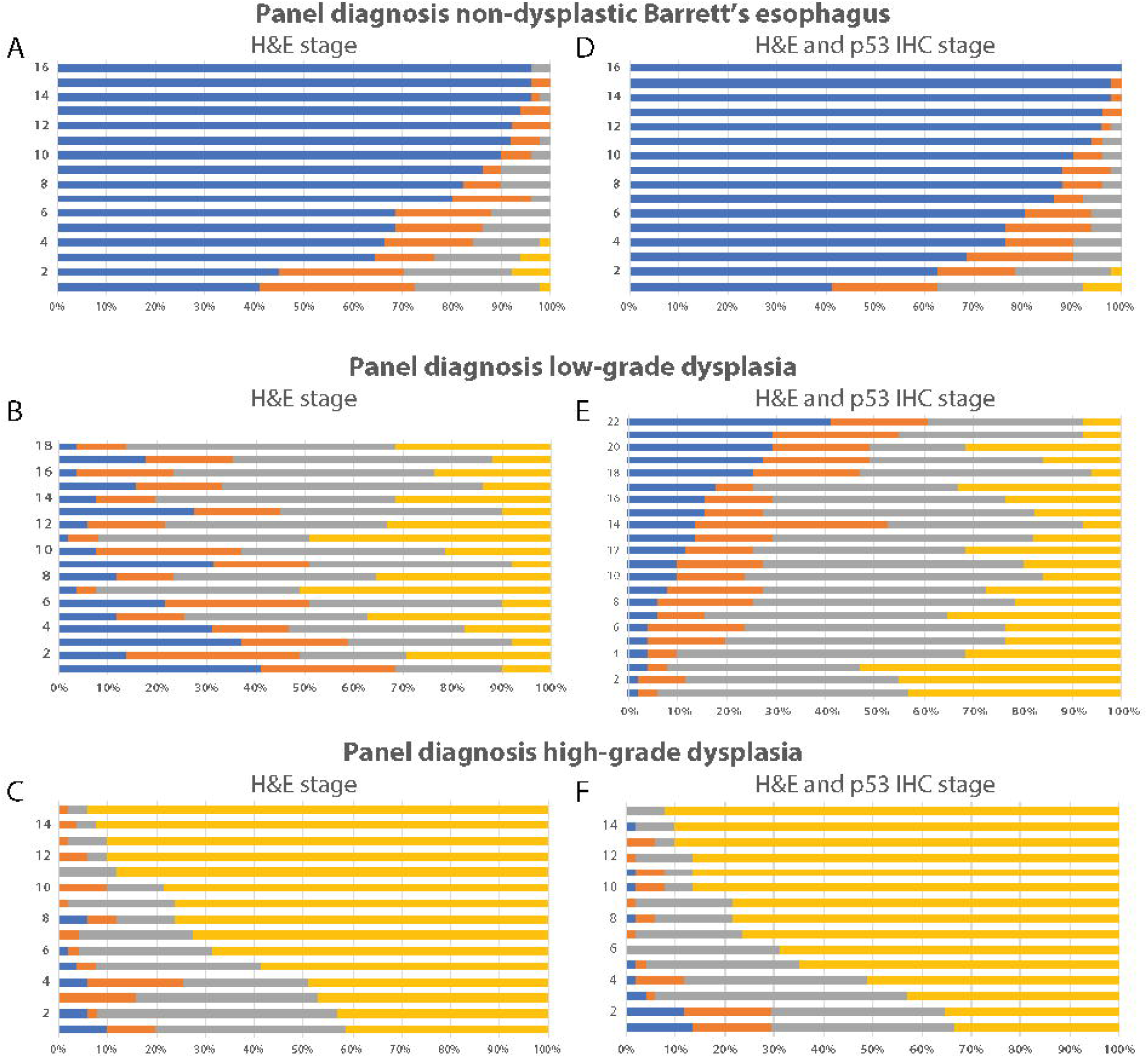
Diagnostic variation per reference diagnoses. A-F) Waterfall plots showing the ranked distribution of case assessments by participating pathologists per diagnostic category, as indicated. Left column (A-C) shows diagnostic variation per reference diagnosis based on H&E slide review alone and right column (D-F) shows diagnostic variation per reference diagnosis after revealing matched p53 IHC. X-axis shows diagnostic concordance in percentages and y-axis shows ranked cases. Color coding as in Figure 2B. Diagnostic variation for indefinite for dysplasia cases is shown in Supplementary Figure 1.

Addition of matched p53 IHC improved diagnostic concordance (**Figure 3D-F** and **Supplementary Figure 1B)** with small but clinically meaningful improvements seen in the diagnostic concordance between participating pathologists for NDBO reference diagnosis cases (83.8% v. 78.8% on H&E slide) and LGD/HGD combined reference diagnosis cases (79.3% v. 77.5% on H&E slide), **Table 2B**. In addition to this, p53 IHC also had a small but beneficial impact on reducing the number of major over and underinterpretations (8.3%, 232 of 2,805 diagnoses), representing 0.5% fewer overall major misinterpretations compared to H&E-stained slide diagnosis alone.

### Characteristics associated with concordance on H&E slides

This massive dataset was then interrogated to search for histopathologist predictors of over or underreporting and major diagnostic errors in univariate analysis. To this end all diagnostic discordances within our dataset (i.e. case diagnoses not matching reference diagnosis) were first reclassified as major or minor over or underinterpretation (see Methods and **Supplementary Table 4**). Factors associated with reduced odds of major diagnostic errors included: ≥5 years of experience commensurate with age (OR 0.65, 95%CI 0.45-0.93); working in an academic teaching hospital (OR 0.59, 95%CI 0.43-0.81); routinely double reporting indefinite for dysplasia cases (OR 0.70, 95%CI 0.52-0.94); working in a larger lab (≥10 versus <10 pathologists OR 0.72, 95%CI 0.54-0.96) and using digital pathology (OR 0.63; 95%CI 0.47-0.89). In contrast, working within a district general hospital (OR 1.72, 95%CI 1.30-2.26) or private hospital (OR 1.41, 95%CI 1.04-1.91), or not using major society guidelines (OR 1.43, 95%CI 1.06-1.94) were all associated with increased odds of major diagnostic errors **(Supplementary Tables 8A-C**).

Several factors were not associated with major diagnostic error, including pathologist sex. Participating in upper gastrointestinal multidisciplinary team meetings was not associated with reduced odds of major diagnostic error, although it was associated with reduced odds of overreporting. Notably, self-identifying as a Barrett’s pathology expert, holding a dedicated fellowship, or reporting greater enjoyment or confidence in Barrett’s reporting were not associated with significant odds of major over or underinterpretation (**Supplementary Table 8A**).

Reporting ≥20 cases per week was associated with reduced odds of over or under-interpretation of Barrett’s dysplasia (OR 0.69, 95%CI 0.53-0.89), although this association was attenuated when investigating major diagnostic errors (**Supplementary Table 8B**).

### Multivariate analyses before and after revealing matched p53 IHC

Multivariable models were then applied, including all factors associated with collective over and underinterpretation on H&E digital slide review in univariate analysis, as shown in **Table 3**. At least 5 years of experience commensurate with age was the strongest protective factor against major diagnostic error on H&E slide review (OR 0.48, 95%CI 0.31-0.74). In contrast, working in a district general hospital was associated with increased odds of major diagnostic error (OR 1.76, 95%CI 1.15-2.69), however this was neutralised if pathologists in these settings viewed cases with additional p53 IHC (OR 1.44, 95%CI 0.92-2.28). As expected, routine use of p53 IHC was associated with reduced odds of major diagnostic error. Viewing 5-19 BO cases with p53 stained slides per week was associated with increased odds of major diagnostic errors, which was neutralised when viewing ≥20 cases per week. Most other results showed similar trends to those seen in univariate analysis, but these were no longer statistically significant (**Table 3**).

**Table 3:** Characteristics associated with odds of major over- or under-interpretation of Barrett’s oesophagus with dysplasia in multivariable adjusted analysis. *All characteristics factors mutually adjusted for each other, **Additional adjustment for p53 immunohistochemical staining in routine pathology practice

| Characteristics* | HE digital slide review |  | p53 IHC stained slide review |  |
| --- | --- | --- | --- | --- |
|  |  | Odds ratio<br>(95%CI) |  | Odds ratio<br>(95% CI)** |
| Age/experience |  |  |  |  |
| Any age/0-4 years experience |  | 1.00 |  | 1.00 |
| Disproportionately more experience to age |  | 0.86 (0.52-1.43) |  | 1.54 (0.89-2.69) |
| 5+ years experience commensurate with age |  | 0.48 (0.31-0.74) |  | 0.89 (0.55-1.45) |
| Interest in Whole slide imaging |  | 0.71 (0.48-1.05) |  | 0.84 (0.56-1.27) |
| Hospital work setting |  |  |  |  |
| Academic teaching hospital |  | 0.96 (0.58-1.60) |  | 1.06 (0.60-1.89) |
| District general hospital |  | 1.76 (1.15-2.69) |  | 1.44 (0.92-2.28) |
| Private hospital |  | 1.09 (0.74-1.62) |  | 0.88 (0.57-1.36) |
| Number Barrett's cases viewed per week |  |  |  |  |
| 0-4 |  | 1.00 |  | 1.00 |
| 5-9 |  | 1.43 (0.92-2.24) |  | 1.77 (1.09-2.89) |
| 10-19 |  | 1.29 (0.82-2.04) |  | 1.71 (1.04-2.80) |
| 20+ |  | 1.55 (0.85-2.81) |  | 0.93 (0.44-1.94) |
| Guidelines used |  |  |  |  |
| North American |  | 1.00 |  | 1.00 |
| British |  | 1.27 (0.77-2.10) |  | 1.04 (0.60-1.78) |
| Japanese |  | 1.23 (0.53-2.85) |  | 0.41 (0.15-1.17) |
| Other |  | 1.40 (0.96-2.05) |  | 1.11 (0.75-1.64) |
| Lab size (number of pathologists) |  |  |  |  |
| <10 |  | 1.00 |  | 1.00 |
| 10+ |  | 0.91 (0.61-1.35) |  | 0.90 (0.60-1.36) |
| Routinely double report indefinite dysplasia |  | 0.78 (0.53-1.14) |  | 1.13 (0.74-1.73) |
| Sometimes routinely stain for p53 |  | Not applicable |  | 0.45 (0.26-0.81) |
| Always/mostly routinely stain for p53 |  | Not applicable |  | 0.30 (0.15-0.60) |
|  | ← Reduced odds 1 Increased odds → |  | ← Reduced odds 1 Increased odds → |  |

## DISCUSSION

We have carried out the largest investigation of diagnostic concordance of BO biopsy reporting amongst gastrointestinal pathologists to date. Previous studies had been limited to a small number of expert pathologists, which meant findings were not necessarily generalizable to real-world settings. This work has revealed several novel findings. First, overall concordance for H&E digital slide review of NDBO and LGD/HGD as a combined outcome was excellent (exceeding 77%), although concordance for IND and LGD as a stand-alone diagnosis was lower (23-42%). These test characteristics replicate known glass slide test characteristics (**Supplementary Table 1**), suggesting that distant BO biopsy slide review is reproducible and safe. Second, the addition of adjunct p53 IHC resulted in small, but clinically meaningful improvements in concordance and a reduction from 8.8% to 8.3% in the prevalence of major misinterpretations of BO dysplasia for pathologists working at a general hospital, signifying that p53 IHC could serve as an extra protection against misdiagnoses for these pathologists working away from teaching hospitals. The limited impact of p53 addition could also be due to a lack of guidelines on staining interpretation. Lastly, multivariate analyses revealed several pathologist characteristics and working practices associated with the prevalence of misinterpretations. Reassuringly, pathologist experience commensurate with age was most protective against major over or underinterpretation, confirming the validity of our experimental strategy. Our multivariate demographic regression analyses also confirm that working within a teaching hospital environment protects against major diagnostic error. This provides supportive evidence for guideline statements that BO complicated by dysplasia is best managed within an expert center. ^21-23 26^.

We found that the overall prevalence of major misinterpretations of dysplastic BO (NDBO classified as LGD/HGD, or vice versa) in this cohort enriched for IND/LGD/HGD cases was 8.8%, which was further reduced, if marginally, by the addition of p53 IHC (8.3%). Although this would suggest a limited impact of the implementation of p53 IHC, our data also reveals that major discordance was reduced by the addition of p53 IHC, specifically for those pathologists working away from teaching hospital settings. Acknowledging that any reduction in the prevalence of major misinterpretation of BO biopsy material is beneficial, these important data suggest that the impact of adjunct p53 IHC is dependent on context and is greatest outside expert center settings. Routine use of p53 IHC labelling is supported by several national guidelines, ^21 23 26^ and our study confirms that this is appropriate.

Taken together, our study for the first time provides an evidence-based quantitative model of BO histopathology diagnosis at expert consensus level. Our findings have several implications for clinical practice of pathology reporting in BO. Diagnostic concordance within a large group of pathologists with different levels of gastrointestinal pathology expertise was excellent for LGD and HGD combined. To implement routine external review of dysplastic BO biopsies, as mandated by several major society guidelines, requires regional teams of dedicated gastrointestinal pathologists. Our data reassuringly suggest that BO reporting on a par with expert consensus is not limited to a small league of experienced histopathologists and can be predicted from a small number of intuitive demographic predictors (experience, professional setting, use of p53 IHC). Combined with our observation that concordance rates for digital slide viewing were not inferior to conventional glass slide pathology review ^18 19^, together these data suggest that distant digital review of problematic BO cases is safe to formally implement within current care delivery systems, provided quality benchmarks are met.

Our study has considerable strengths compared to previous interobserver variation studies of BO reporting. We have evaluated diagnostic concordance for dysplastic BO amongst the largest group of gastrointestinal pathologists worldwide. The heterogeneous mix of pathologists involved in this study also enabled novel investigations into pathologist-dependent predictors associated with diagnostic discordance. The online reporting strategy mimicked routine workflow and facilitated data collection and curation in a flexible manner. The case set was purposely enriched for dysplastic cases in order to attain sufficient statistical power in our downstream regression analyses.

This study also has limitations that are important to note. First, acknowledging that the diagnostic spectrum of BO represents a morphologic continuum, we aimed to include diagnostic categories from across the diagnostic spectrum, including arbitrary cases that straddle the transition from low-grade dysplasia to high-grade dysplasia, as well as indefinite for dysplasia cases. Our case-mix for this reason does not represent a cross-section of diagnostic biopsy cases encountered in daily practice, which would be heavily weighted towards the NDBO end of the spectrum. A second limitation is that while our heterogeneous global group of pathologists allowed us to interrogate associations of a host of operator-dependent characteristics with diagnostic consensus (case volume, practice setting, diagnostic experience, etc.), this study feature may limit the generalizability of our findings within the national setting. Replication of our findings in samples of pathologists within particular geographic regions adhering to one diagnostic guideline will be required to determine whether the quantitative predictive features described here are similarly applicable in that setting. Given that the majority of pathologists participating in this study were based either in Europe or North America, greater representation from low to middle income settings would be particularly welcome. This could further enhance the value of this recursive exercise for teaching and registration purposes.

In conclusion, using this rich dataset of case assessments by a large, heterogeneous sample of gastrointestinal pathologists, we have evaluated diagnostic concordance for BO diagnosis using digital case review. Our results reveal quantitative predictors of diagnostic performance that will aid formulation of quality assurance criteria for guideline development and standard implementation of digital pathology in BO biopsy review.

## Data Availability

All primary data will be made available upon request

## LIST OF ABBREVIATIONS

BO: Barrett’s oesophagus
BMI: body mass index
CI: confidence interval
CRF: case record form
OAC: oesophageal adenocarcinoma
HGD: high-grade dysplasia
IHC: immunohistochemistry
IMC: intramucosal carcinoma
IND: indefinite for dysplasia
IQR: interquartile range
K: kappa value
LGD: low-grade dysplasia
NDBO: non-dysplastic Barrett’s oesophagus
OR: odd’s ratio
WSI: whole slide imaging

## ACKNOWLEDGEMENTS

We sincerely thank Joann Elmore and Gary Longdon for their helpful additions to our study protocol. We sincerely thank Alden van Putten, Rudy Scholten, Rene Breet and David de Koning (Open Clinica) for their help in building and maintaining the on-line study environment. We sincerely thank Onno de Boer, Eelco Roos, and Wim van Est for their help scanning of the slides.

---

**# BOLERO STUDY PARTICIPANTS (in alphabetical order)**

Dr. Junko Aida, Tokyo Metropolitan Institute of Gerontology, Tokyo, Japan

Dr. Rossana Baiocco, General Hospital of Desenzano del Garda, Desenzano, Italy

Dr. Camille Boulagnon-Rombi, Université de Reims Champagne-Ardenne, Reims, France

Dr. Iva Brcic, Medical University of Graz, Graz, Austria

Dr. Lodewijk Brosens, University Medical Center Utrecht, Utrecht, the Netherlands

Dr. Fátima Carneiro, IPATIMUP, Porto, Portugal

Dr. Gieri Cathomas, Kantosspital Baselland, Liestal, Switzerland

Dr. Denis Chatelain, CHU Amiens-Picardie, Amiens, France

Dr. Allison Cluroe, Addenbrookes Hospital, Cambridge, United Kingdom

Dr. Parag Dabir, Regional Hospital, Randers, Denmark

Dr. Giovanni De Petris, Penrose Hospital, Colorado Springs, United States of America

Dr. Michael Doukas, Erasmus Medical Center, Rotterdam, the Netherlands

Dr. Hala El-Zimaity, Toronto General Hospital, Toronto, Canada

Dr. Matteo Fassan, University of Padua, Padua, Italy

Dr. Roberto Fiocca, University of Genova, Genova, Italy

Dr. Jean-François Fléjou, Saint Antoine Hospital, Paris, France

Dr. Alejandro García Varona, Hospital El Bierzo, Leon, Spain

Dr. Elvira Gonzalez Obeso, Hospital Clinico Universitario, Valladolid, Spain

Dr. Heike Grabsch, 1. Division of Pathology and Data Analytics, Leeds Institute of Medical Research at St James’s, University of Leeds, Leeds, UK, 2. Department of Pathology, GROW School for Oncology and Developmental Biology, Maastricht University Medical Center+, Maastricht, NL

Dr. Federica Grillo, University of Genova, Genova, Italy

Dr. Barbara Gruber, Patologia Bariloche, San Carlos de Bariloche, Argentina

Dr. Laura Guerra Pastrian, University Hospital La Paz, Madrid, Spain

Dr. Anne Hoorens, University Hospital Gent, Gent, Belgium

Dr. Marnix Jansen, University College Hospital, London, United Kingdom

Dr. Katerina Kamaradova, Charles University Hospital, Hradec Kralove, Czech Republic

Dr. Ryoji Kushima, Shiga University of Medical Science, Shiga, Japan

Dr. Cord Langner, Medical University of Graz, Graz, Austria

Dr. Rupert Langer, University of Bern, Bern, Switzerland

Dr. Felix Lasitschka, Universitätsklinikum Heidelberg, Heidelberg, Germany

Dr. Ester Lörinc, University Hospital Lund and Malmö, Lund, Sweden

Dr. Luca Mastracci, University of Genova, Genova, Italy

Dr. Damian McManus, Belfast HSC Trust, Belfast, Northern Ireland

Dr. Sybren Meijer, Academic Medical Center Amsterdam, the Netherlands

Dr. Carmen Mendez, University Hospital La Paz, Madrid, Spain

Dr. Anya Milne, Diakonessenhuis, Utrecht, the Netherlands

Dr. Miriam Mitchison, University College Hospital London, United Kingdom

Dr. Masoud Mireskandari, Jena University Hospital, Jena, Germany

Dr. Elizabeth Montgomery, Johns Hopkins Medical Institute, Baltimore, United States of America

Dr. Cian Muldoon, St. James’s Hospital, Dublin, Ireland

Dr. Maria O’Donovan, Cambridge Cancer Centre, Cambridge, United Kingdom

Dr. Rob Odze, Brigham and Women’s Hospital, Boston, United States of America

Dr. Johan Offerhaus, University Medical Center Utrecht, the Netherlands

Dr. Gabriel Olmedilla, University Hospital La Paz, Madrid, Spain

Dr. John Pauli, The Prince Charles Hospital, Brisbane, Australia

Dr. Rachel S. van der Post, Radboud university medical centre, Nijmegen, the Netherlands

Dr. Bob Riddell, Mount Sinai Hospital, Toronto, Canada

Dr. Ari Ristimaki, Haartman Institute, Helsinki, Finland

Dr. Ana Rodriguez, University Hospital La Paz, Madrid, Spain

Dr. Manual Rodriguez-Justo, University College Hospital, London, United Kingdom

Dr. Shigeki Sekine, National Cancer Center Hospital, Tokyo, Japan

Dr. Kees Seldenrijk, St. Antonius Hospital, Nieuwegein, the Netherlands

Dr. Tulio Souza, Hospital Aliança, Salvador, Brazil

Dr. Matt Stachler, Brigham and Women’s Hospital, Boston, United States of America

Dr. Michael Vieth, Klinikum Bayreuth, Bayreuth, Germany

Dr. Vincenzo Villanacci, Spedali Civili di Brescia, Brescia, Italy

Dr. Rhonda Yantiss, Weill Cornell Medical College, New York, United States of America

**Supplementary Figure 1:**
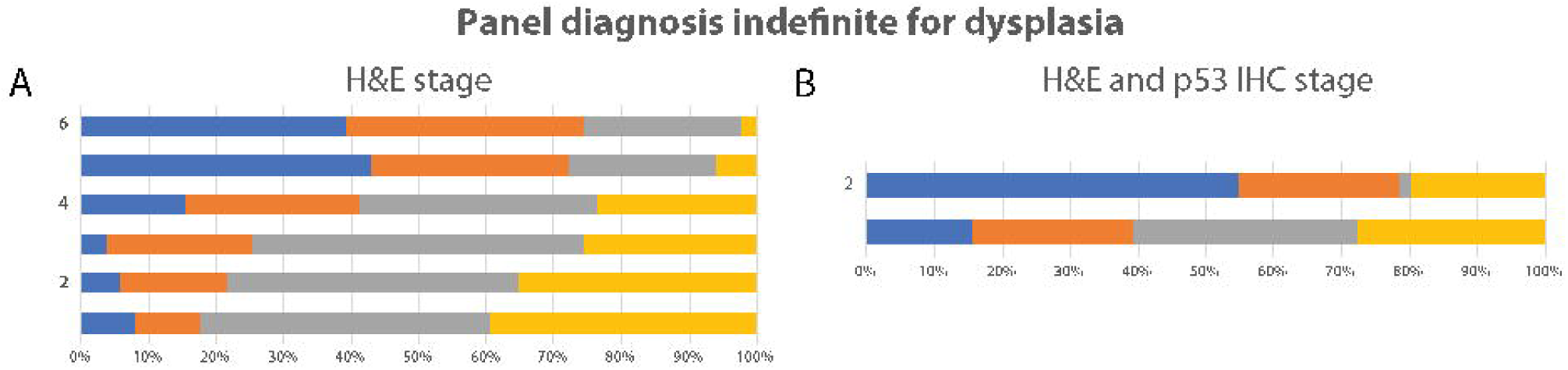
Diagnostic variation for indefinite for dysplasia diagnoses before (A) and after (B) revealing matched p53 IHC labelling. X-axis shows diagnostic concordance in percentages and y-axis shows ranked cases. See text for details.

**Supplementary Table 1:** Overview concordance studies in Barrett’s oesophagus. *representing interobserver agreement unless mentioned otherwise

| Author | Year | Journal | No of cases | No of review pathologists | No of rounds | Group discussion | Use of p53 IHC | Type of observer agreement | K* total | K* NDBO | K* LGD | K* HGD / IMC | K* IND |
| --- | --- | --- | --- | --- | --- | --- | --- | --- | --- | --- | --- | --- | --- |
| Coco (4) | 2011 | Am J Surg Pathol | Set 1: 40, Set 2: 63 | 6 | 1 per set | Yes (between sets) | No | Interobserver | Set 1: 0.44<br>set 2: 0.47 | Set 1: 0.57, Set 2: 0.50 | Set 1: 0.31, Set 2: 0.40 | Set 1: 0.67, Set 2: 0.72 | Set 1: 0.018, Set 2: 0.014 |
| Horvath (5) | 2014 | J Gastroent & Hep | 85 | 6 | 1 | No | No | Interobserver (Fleiss) | 0.33 | - | - | - | - |
| Kaye (6) | 2009 | Histopathol | 186 | 5 | 2 | Yes | Yes | Interobserver (weighted pairs) | Without p53 IHC*: 0.5-0.65<br>With p53 IHC*: 0.53-0.70 |  |  |  |  |
| Kaye (7) | 2016 | Histopathol | 72 | 10 | 2 | Yes (before sets) | Yes | Interobserver (weighted pairs) | Without p53 IHC: 0.47<br>With p53 IHC*: 0.55 |  |  |  |  |
| Kerkhof (8) | 2007 | Histopathol | 793 | 11 | 1 | Yes (in case of discrepancies) | No | Interobserver (unweighted) | 0.25 | 0.27 | - | 0.58 |  |
| Lim (9) | 2007 | Endoscopy | 88 | 5 | 1 | No | No | Interobserver | 0.48 (range 0.42-0.70) | - | - | - | - |
| Montgomery (10) | 2001 | Human Path | 250 | 12 | 2 | Yes (between 2 sets) | No | Intraobserver / Interobserver | Intraobserver: 0.60<br>Interobserver: 0.43 |  |  |  |  |

| Author | Year | Journal | No of cases | No of review pathologists | No of rounds | Group discussion | Use of p53 IHC | Type of observer agreement | K* total | K*NDBO | K* LGD | K* HGD / IMC | K* IND |
| --- | --- | --- | --- | --- | --- | --- | --- | --- | --- | --- | --- | --- | --- |
| Pech (11) | 2007 | Scand J Gastroenterol | 50 | 2 | 1 | No | No | Interobserver (unweighted) |  |  | 0.69 (2 experts)<br>0.03 (2 experts vs general pathologists) |  |  |
| Sanders (12) | 2012 | Histopathol | 61 | 5 | 2 | No | Yes | Interobserver | R1: 0.71 (Fleiss)<br>R1 for subgroup: 0.60 (conventional microscopy)<br>R2: 0.44 (digital microscopy) |  |  |  |  |
| Sangle (13) | 2015 | Modern Path | 437 | 3 | 2 | No | Yes | Interobserver |  |  |  | 0.77 |  |
| Skacel (14) | 2000 | AJG | 100 | 3 | 1 | No | Unknown | Interobserver (unweighted) |  |  | 0.17 (mean) |  |  |
| Skacel (15) | 2002 | AJG | 16 | 3 | 1 | No | Yes | Sensitivity / specificity |  |  | With p53 IHC*: sensitivity 100%, specificity 75% |  |  |
| Sonwalkar (16) | 2010 | Histopathol | 101 | 3 | 1 | No | No | Interobserver (weighted) | 0.35 | 0.73 | 0.29 | 0.43 | 0.18 |
| Wani (17) | 2011 | Gastroenterol | 88 | 2 | 1 | No | No | Interobserver (unweighted) |  |  | 0.14 |  |  |

**Supplementary Table 2:** Demographic and Clinical Characteristics of patient biopsies.

| **Characteristics** | **Number of patients**<br>**n=55 (%)** |
| --- | --- |
| Male | 52 (94.5) |
| Age, years (median, range) | 65 (36-86) |
| BMI*, kg/m<sup>2</sup>, median (IQR) | 27 (3.9) |
| History of smoking | 35 (63.6) |
| If so, mean number of pack years | 14 |
| Heart burn symptoms | 49 (89.1) |
| Anti-reflux medication | 53 (96.4) |
| Circumferential Barrett's extent, cm, median (IQR) | 4 (7.8) |
| Length of Barrett's segment, cm, median (IQR) | 5 (8) |
| Consensus diagnoses on H&E slide, before p53 IHC |  |
| NDBO | 16 (29.1) |
| IND | 6 (10.9) |
| LGD | 18 (32.7) |
| HGD | 15 (27.3) |
| Consensus diagnosis, after p53 IHC |  |
| NDBO | 16 (29.1) |
| IND | 2 (3.6) |
| LGD | 22 (40.0) |
| HGD | 15 (27.3) |

**Supplementary Table 3:** Demographic questionnaire. *free text field

| **Question** | **Answer options** |
| --- | --- |
| **Part 1: General demographic information** |  |
| Your age | 30-39 / 40-49 / 50-59 / 60 or above |
| Your gender | Male / Female |
| Do you work in an academic teaching hospital? | Yes / No |
| Do you work in a district general hospital? | Yes / No |
| Do you work in a private practice? | Yes / No |
| Did you participate in a GI-pathology fellowship? | Yes / No |
| **Part 2: Professional Experience** |  |
| Your practice size: | <10 pathologists / 10 pathologists or more |
| Years' experience in signing out Barrett's biopsy cases: | 0-4 / 5-9 / 10-19 / 20 or more |
| Which guidelines do you adhere to in sign-out practice of Barrett's esophagus? | • North-American (ACG) Guidelines (*Shaheen et al. AJG 2016*)<br>• British (BSG) Guidelines (*Fitzgerald et al. Gut 2013*)<br>• Guidelines Japanese Society for Esophageal Diseases (*Kuwano et al. Esophagus 2012*)<br>• Cancer Council Australia Guidelines (*Whiteman et al. JGH 2015*)<br>• Other |
| Total no. of Barrett's biopsy cases reviewed per week (including local, referral, surveillance, and new diagnoses): | 0-4 / 5-9 / 10-19 / 20-29 / 30-39 / 40 or more / don't know |
| Within your team of consultants, are you the designated local expert for complicated Barrett's biopsy cases? | Yes / No / don't know |
| Do you generally feel confident when signing out Barrett's dysplasia specimens? | Scale of 1-6, where 1=very confident and 6=not confident |
| Do you enjoy signing out Barrett's specimens? | Scale of 1-6 where 1=very much and 6=not at all |
| Do you also sign out endoscopic mucosal resection (EMR) specimens? | Yes / No |
| If Yes: On average, how many EMR specimens do you sign out on a weekly basis? | <1 / 1 / 2-5 / 6-10 / 11-20 / >20 |
| Do you receive an endoscopy report with most esophageal biopsy series and/or EMR cases? | Yes / No |
| If Yes: Do you feel the endoscopy report generally provides you with enough information to answer the clinical request? | Yes / No |
| In your experience, do endoscopists in your institution generally adhere to the Seattle surveillance protocol (quadratic biopsies every 2 | Always / Most of the time / Some of the time / Never |
| cm taken in separate containers)? |  |
| Are target biopsies of nodules and other suspicious areas sent in separate containers? | Always / Most of the time / Some of the time / Never |
| Do you IHC label for p53 on Barrett's surveillance biopsies? | Always / Most of the time / Some of the time / Never |
| Are Barrett's dysplasia or indefinite for dysplasia cases routinely double reported? | Yes / No |
| Do you take part in regular upper gastrointestinal multidisciplinary meetings? | Yes / No |
| **Part 3: Experience with digital pathology** |  |
| Does your laboratory make use of whole slide imaging (digital pathology)? | Yes / No / Don't know |
| If Yes: type of use: | Research purposes / External consultation and consensus panels / Digitalized laboratory / Other; namely....* |
| Are you interested in digital pathology? | Scale of 1-6 where 1=very interested and 6=not interested |
| Do you think digital pathology can completely replace light microscopy? | Yes / No |

**Supplementary Table 4:** Overview of diagnostic errors classification.

| **Participating pathologist diagnosis** | **Reference panel pathologists' diagnosis** | **Diagnostic class** | **Number of cases on HE staining / on HE and p53 IHC staining** |
| --- | --- | --- | --- |
| LGD | NDBO | Major overinterpretation | 151/153 |
| HGD | NDBO | Major overinterpretation | 17/21 |
| IND | NDBO | Minor overinterpretation | 59/36 |
| LGD | IND | Minor overinterpretation | 165/178 |
| HGD | IND | Minor overinterpretation | 45/38 |
| HGD | LGD | Minor overinterpretation | 159/165 |
| NDBO | LGD | Major underinterpretation | 71/53 |
| NDBO | HGD | Major underinterpretation | 9/5 |
| NDBO | IND | Minor underinterpretation | 93/74 |
| IND | LGD | Minor underinterpretation | 110/27 |
| IND | HGD | Minor underinterpretation | 67/15 |
| LGD | HGD | Minor underinterpretation | 220/275 |

**Supplementary Table 5:** Odds ratios for the association with major over or underinterpretation*. *According to mutually adjusted regression models for age and experience. This information was used to generate three categories of age/experience combinations used in further multivariable-adjusted models: green; category 1: Pathologists with 0-4 years experience, regardless of age (Reference category), orange; category 2: Pathologists with disproportionately greater years of experience relative to age (combined OR 1.36, 95% CI 0.90-2.60), blue; category 3: Pathologists with experience commensurate with age (combined OR 0.65, 95% CI 0.45-0.93), **Significant result (OR 0.47, 95% CI 0.28-0.78). All other results not significant.

|  |  | Experience (yrs) |  |  |  |
| --- | --- | --- | --- | --- | --- |
|  |  | 0-4 | 5-9 | 10-19 | 20+ |
| Age (yrs) | 30-40 | Reference | 1.40 | 1.24 | N/A |
|  | 41-50 | 1.04 | 0.69 | 0.47** | 1.39 |
|  | 51-60 | N/A | 0.57 | 0.64 | 0.86 |
|  | 60+ | N/A | N/A | N/A | 0.75 |

**Supplementary Table 6:**
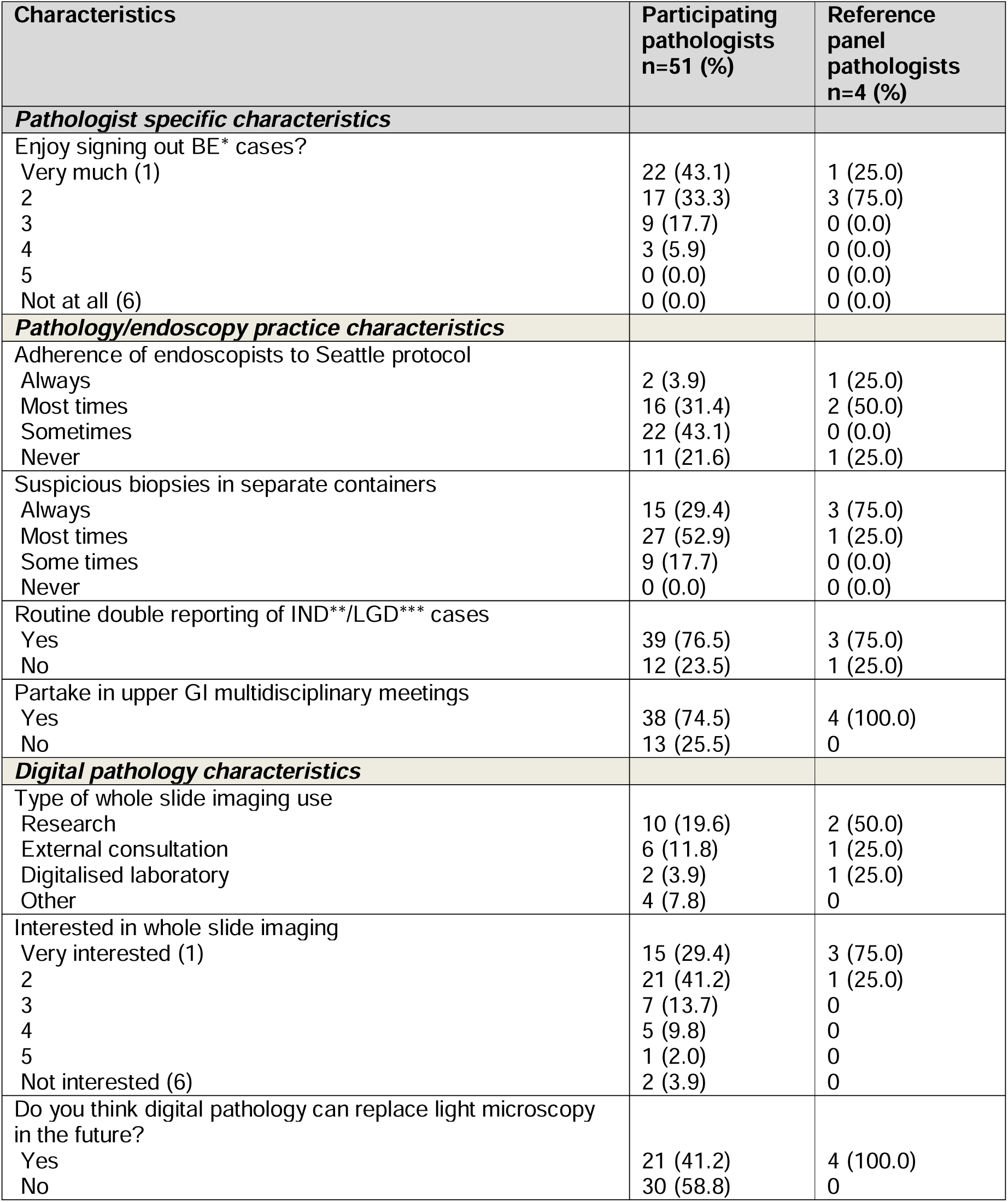
Demographics of pathologists reporting in the BOLERO study (continued).

| Characteristics | Participating pathologists<br>n=51 (%) | Reference panel pathologists<br>n=4 (%) |
| --- | --- | --- |
| ***Pathologist specific characteristics*** |  |  |
| Enjoy signing out BE* cases? |  |  |
| Very much (1) | 22 (43.1) | 1 (25.0) |
| 2 | 17 (33.3) | 3 (75.0) |
| 3 | 9 (17.7) | 0 (0.0) |
| 4 | 3 (5.9) | 0 (0.0) |
| 5 | 0 (0.0) | 0 (0.0) |
| Not at all (6) | 0 (0.0) | 0 (0.0) |
| ***Pathology/endoscopy practice characteristics*** |  |  |
| Adherence of endoscopists to Seattle protocol |  |  |
| Always | 2 (3.9) | 1 (25.0) |
| Most times | 16 (31.4) | 2 (50.0) |
| Sometimes | 22 (43.1) | 0 (0.0) |
| Never | 11 (21.6) | 1 (25.0) |
| Suspicious biopsies in separate containers |  |  |
| Always | 15 (29.4) | 3 (75.0) |
| Most times | 27 (52.9) | 1 (25.0) |
| Some times | 9 (17.7) | 0 (0.0) |
| Never | 0 (0.0) | 0 (0.0) |
| Routine double reporting of IND**/LGD*** cases |  |  |
| Yes | 39 (76.5) | 3 (75.0) |
| No | 12 (23.5) | 1 (25.0) |
| Partake in upper GI multidisciplinary meetings |  |  |
| Yes | 38 (74.5) | 4 (100.0) |
| No | 13 (25.5) | 0 |
| ***Digital pathology characteristics*** |  |  |
| Type of whole slide imaging use |  |  |
| Research | 10 (19.6) | 2 (50.0) |
| External consultation | 6 (11.8) | 1 (25.0) |
| Digitalised laboratory | 2 (3.9) | 1 (25.0) |
| Other | 4 (7.8) | 0 |
| Interested in whole slide imaging |  |  |
| Very interested (1) | 15 (29.4) | 3 (75.0) |
| 2 | 21 (41.2) | 1 (25.0) |
| 3 | 7 (13.7) | 0 |
| 4 | 5 (9.8) | 0 |
| 5 | 1 (2.0) | 0 |
| Not interested (6) | 2 (3.9) | 0 |
| Do you think digital pathology can replace light microscopy in the future? |  |  |
| Yes | 21 (41.2) | 4 (100.0) |
| No | 30 (58.8) | 0 |

**Supplementary Table 7:** Cross table comparing the 4 reference pathologist diagnoses to the consensus-derived reference diagnoses for 55 esophageal biopsy cases (a) on HE staining and (b) on HE and p53 IHC staining for 440 total case interpretations*. *Overall concordance for 154/220 diagnoses (70%, 95%CI 63.9-76.1%), increasing to 178/220 (80.9%, 95%CI 75.7-86.1%) when LGD and HGD were combined, **Note consensus reference panel results are scaled x4 to allow for comparison versus the four individual panel members, who contributed to the consensus reference panel, preconsensus results. Results represent 220 diagnoses in 55 oesophageal biopsy cases. ***Overall concordance for 169/220 diagnoses (76.8%, 95%CI 71.2-82.4%), increasing to 195/220 (88.6%, 95%CI 84.4-92.8%) when LGD and HGD were combined.

|  | Consensus<br>reference<br>panel** | Reference panel members'<br>individual diagnoses<br>(preconsensus) |  |  |  | % Concordance |  |  |
| --- | --- | --- | --- | --- | --- | --- | --- | --- |
|  |  |  |  |  |  | Under-<br>interpret | Over-<br>interpret | Concorda<br>nce |
| a. Before addition of p53 immunohistochemistry |  |  |  |  |  |  |  |  |
| Diagnosis |  | ND | IND | LGD | HGD |  |  |  |
| NDBO | 64 | 54 | 9 | 1 | 0 | / | 15.6<br>(6.7-24.5) | 84.4<br>(75.5-93.3) |
| IND | 24 | 7 | 6 | 9 | 2 | 29.2<br>(10.0-48.4) | 45.8<br>(24.7-66.9) | 25<br>(6.7-43.3) |
| LGD | 72 | 3 | 10 | 47 | 12 | 18.0<br>(9.1-26.9) | 16.7<br>(8.1-25.3) | 65.3<br>(54.3-76.3) |
| HGD | 60 | 0 | 1 | 12 | 47 | 21.7<br>(11.3-32.1) | / | 78.3<br>(67.9-88.7) |
| LGD or HGD | 132 | 3 | 11 | 118 |  | 10.6<br>(5.3-15.9) | / | 89.4<br>(84.1-94.7) |
| Total | 220 |  |  |  |  |  |  |  |
|  | Consensus<br>reference<br>panel*** | Reference panel members'<br>individual diagnoses<br>(preconsensus) |  |  |  | % Concordance |  |  |
|  |  |  |  |  |  | Under-<br>interpret | Over-<br>interpret | Concorda<br>nce |
| b. After addition of p53 immunohistochemistry |  |  |  |  |  |  |  |  |
| Diagnosis |  | ND | IND | LGD | HGD |  |  |  |
| ND | 64 | 55 | 6 | 3 | 0 | / | 14.1<br>(5.6-22.6) | 85.9<br>(77.4-94.4) |
| IND | 8 | 2 | 4 | 1 | 1 | 25<br>(0-61.1) | 25<br>(0-61.1) | 50<br>(8.3-91.7) |
| LGD | 88 | 4 | 7 | 64 | 13 | 12.5<br>(5.6-19.4) | 14.8<br>(7.4-22.2) | 72.7<br>(63.4-82.0) |
| HGD | 60 | 0 | 1 | 13 | 46 | 23.3<br>(12.6-34.0) | / | 76.7<br>(66.0-87.4) |
| LGD or HGD | 148 | 4 | 8 | 136 |  | 8.1<br>(3.7-12.5) | / | 91.9<br>(87.5-96.3) |
| Total | 220 |  |  |  |  |  |  |  |

**Supplementary table 8:** (a) Individual pathologist features and odds of over or underreporting Barrett’s dysplasia: unadjusted analysis.

| Variable | No. correct diagnoses | No. Over-reported diagnoses | Overreporting OR (95% CI) | No. Under-reported diagnoses | Underreporting OR (95% CI) | Over or Underreporting OR (95% CI) | No. Major over- or under-reported diagnoses | Major over- or Underreporting OR (95% CI) |
| --- | --- | --- | --- | --- | --- | --- | --- | --- |
| **Total numbers** | **n=1639** | **n=570** |  | **n=596** |  |  | **n=582** |  |
| Age, years |  |  |  |  |  |  |  |  |
| 30-39 | 393 | 159 | 1.00 | 163 | 1.00 | 1.00 | 86 | 1.00 |
| 40-49 | 576 | 182 | 0.78 (0.61-1.00) | 177 | **0.74 (0.58-0.95)** | **0.76 (0.62-0.93)** | 70 | **0.56 (0.40-0.78)** |
| 50-59 | 452 | 138 | **0.75 (0.58-0.98)** | 180 | 0.96 (0.75-1.23) | 0.86 (0.70-1.05) | 62 | **0.63 (0.44-0.89)** |
| 60+ | 218 | 91 | 1.03 (0.76-1.40) | 76 | 0.84 (0.61-1.16) | 0.93 (0.73-1.20) | 30 | **0.63 (0.40-0.98)** |
| Experience, years |  |  |  |  |  |  |  |  |
| 0-4 | 249 | 123 | 1.00 | 68 | 1.00 | 1.00 | 46 | 1.00 |
| 5-9 | 268 | 98 | 0.74 (0.54-1.02) | 129 | **1.76 (1.25-2.48)** | 1.10 (0.85-1.43) | 56 | 1.13 (0.74-1.73) |
| 10-19 | 609 | 204 | **0.68 (0.52-0.89)** | 177 | 1.06 (0.78-1.46) | 0.82 (0.65-1.02) | 63 | **0.56 (0.37-0.84)** |
| 20+ | 513 | 145 | **0.57 (0.43-0.76)** | 222 | **1.59 (1.16-2.16)** | 0.93 (0.74-1.18) | 83 | 0.88 (0.59-1.29) |
| Age/experience combination |  |  |  |  |  |  |  |  |
| 0-4 years exp./All ages | 249 | 123 | 1.00 | 68 | 1.00 | 1.00 | 46 | 1.00 |
| Disproportionate more exp/age | 274 | 77 | **0.57 (0.41-0.79)** | 144 | **1.92 (1.38-2.69)** | 1.05 (0.81-1.36) | 69 | 1.36 (0.90-2.06) |
| Exp. commensurate with age | 1116 | 370 | **0.67 (0.51-0.86)** | 384 | 1.26 (0.94-1.69) | 0.88 (0.71-1.09) | 133 | **0.65 (0.45-0.93)** |
| Sex |  |  |  |  |  |  |  |  |
| Male | 936 | 333 | 1.00 | 326 | 1.00 | 1.00 | 145 | 1.00 |
| Female | 703 | 237 | 0.95 (0.78-1.15) | 270 | 1.10 (0.91-1.33) | 1.02 (0.88-1.19) | 103 | 0.95 (0.72-1.24) |
| Fellowship |  |  |  |  |  |  |  |  |
| No | 750 | 284 | 1.00 | 231 | 1.00 | 1.00 | 102 | 1.00 |
| Yes | 889 | 286 | 0.85 (0.70-1.03) | 365 | **1.33 (1.10-1.61)** | 1.07 (0.92-1.24) | 146 | 1.21 (0.92-1.58) |
| Barrett's expert? |  |  |  |  |  |  |  |  |
| No | 257 | 108 | 1.00 | 75 | 1.00 | 1.00 | 36 | 1.00 |
| Yes | 1098 | 357 | 0.77 (0.60-1.00) | 415 | 1.30 (0.98-1.72) | 0.99 (0.80-1.22) | 160 | 1.04 (0.71-1.53) |
| Don't Know | 284 | 105 | 0.88 (0.64-1.21) | 106 | 1.28 (0.91-1.80) | 1.04 (0.80-1.35) | 52 | 1.31 (0.83-2.06) |
| Confidence |  |  |  |  |  |  |  |  |
| 3/4 (moderate) | 489 | 202 | 1.00 | 189 | 1.00 | 1.00 | 84 | 1.00 |
| 1/2 (very) | 1150 | 368 | **0.78 (0.63-0.95)** | 407 | 0.92 (0.75-1.12) | 0.84 (0.72-0.99) | 164 | 0.83 (0.63-1.10) |
| Enjoy |  |  |  |  |  |  |  |  |
| 3/4 (moderate) | 371 | 131 | 1.00 | 158 | 1.00 | 1.00 | 60 | 1.00 |
| 1/2 (very) | 1268 | 439 | 0.98 (0.78-1.23) | 438 | 0.81 (0.65-1.01) | 0.89 (0.74-1.06) | 188 | 0.92 (0.67-1.25) |

**Supplementary table 8:**
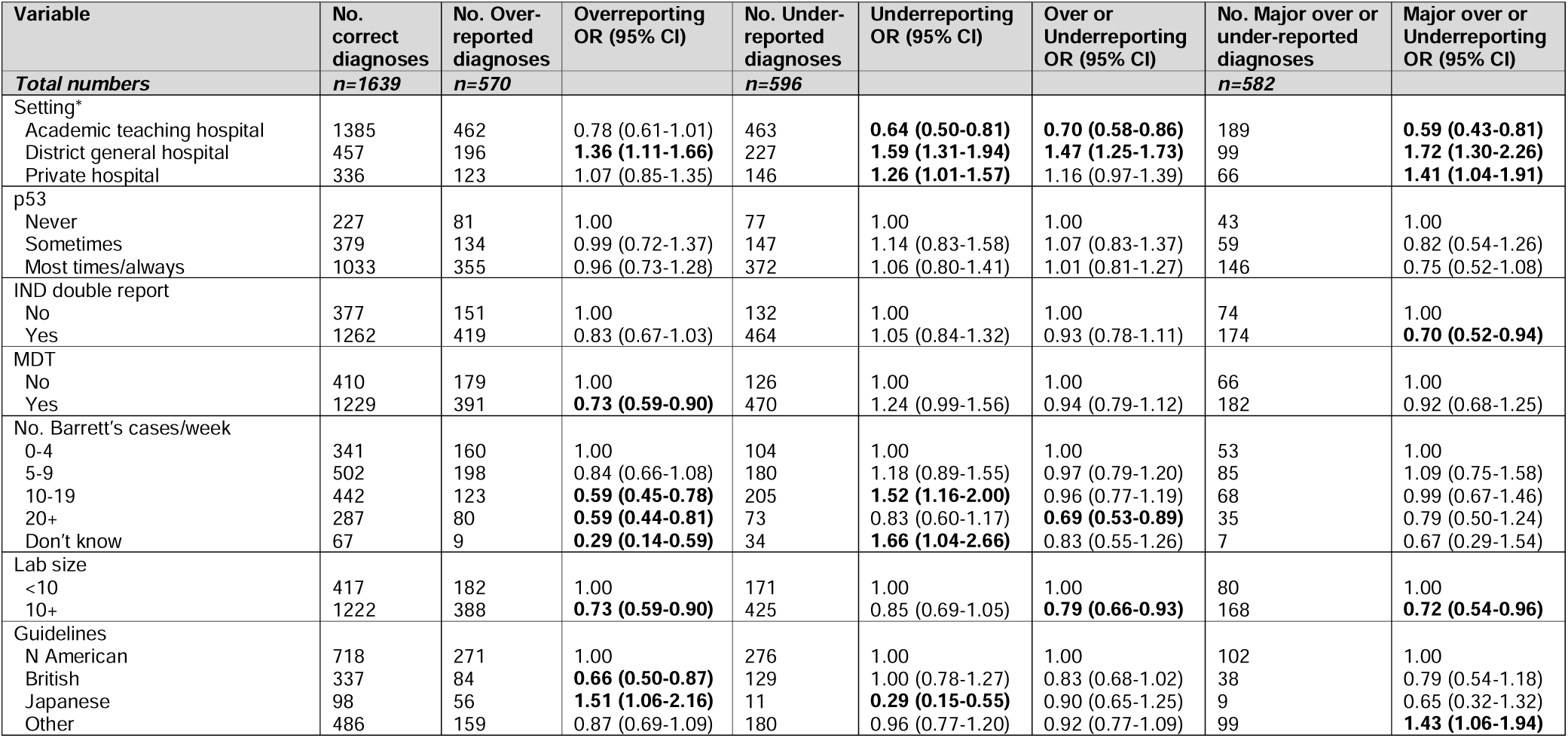
(b) Pathologist working practices and odds of over or underreporting Barrett’s dysplasia: unadjusted analysis. *Reference is not working within these settings. Some pathologists work in multiple settings.

| Variable | No. correct diagnoses<br>*n*=1639 | No. Over-reported diagnoses<br>*n*=570 | Overreporting OR (95% CI) | No. Under-reported diagnoses<br>*n*=596 | Underreporting OR (95% CI) | Over or Underreporting OR (95% CI) | No. Major over or under-reported diagnoses<br>*n*=582 | Major over or Underreporting OR (95% CI) |
| --- | --- | --- | --- | --- | --- | --- | --- | --- |
| **Total numbers** |  |  |  |  |  |  |  |  |
| Setting* |  |  |  |  |  |  |  |  |
| Academic teaching hospital | 1385 | 462 | 0.78 (0.61-1.01) | 463 | **0.64 (0.50-0.81)** | **0.70 (0.58-0.86)** | 189 | **0.59 (0.43-0.81)** |
| District general hospital | 457 | 196 | **1.36 (1.11-1.66)** | 227 | **1.59 (1.31-1.94)** | **1.47 (1.25-1.73)** | 99 | **1.72 (1.30-2.26)** |
| Private hospital | 336 | 123 | 1.07 (0.85-1.35) | 146 | **1.26 (1.01-1.57)** | 1.16 (0.97-1.39) | 66 | **1.41 (1.04-1.91)** |
| p53 |  |  |  |  |  |  |  |  |
| Never | 227 | 81 | 1.00 | 77 | 1.00 | 1.00 | 43 | 1.00 |
| Sometimes | 379 | 134 | 0.99 (0.72-1.37) | 147 | 1.14 (0.83-1.58) | 1.07 (0.83-1.37) | 59 | 0.82 (0.54-1.26) |
| Most times/always | 1033 | 355 | 0.96 (0.73-1.28) | 372 | 1.06 (0.80-1.41) | 1.01 (0.81-1.27) | 146 | 0.75 (0.52-1.08) |
| IND double report |  |  |  |  |  |  |  |  |
| No | 377 | 151 | 1.00 | 132 | 1.00 | 1.00 | 74 | 1.00 |
| Yes | 1262 | 419 | 0.83 (0.67-1.03) | 464 | 1.05 (0.84-1.32) | 0.93 (0.78-1.11) | 174 | **0.70 (0.52-0.94)** |
| MDT |  |  |  |  |  |  |  |  |
| No | 410 | 179 | 1.00 | 126 | 1.00 | 1.00 | 66 | 1.00 |
| Yes | 1229 | 391 | **0.73 (0.59-0.90)** | 470 | 1.24 (0.99-1.56) | 0.94 (0.79-1.12) | 182 | 0.92 (0.68-1.25) |
| No. Barrett's cases/week |  |  |  |  |  |  |  |  |
| 0-4 | 341 | 160 | 1.00 | 104 | 1.00 | 1.00 | 53 | 1.00 |
| 5-9 | 502 | 198 | 0.84 (0.66-1.08) | 180 | 1.18 (0.89-1.55) | 0.97 (0.79-1.20) | 85 | 1.09 (0.75-1.58) |
| 10-19 | 442 | 123 | **0.59 (0.45-0.78)** | 205 | **1.52 (1.16-2.00)** | 0.96 (0.77-1.19) | 68 | 0.99 (0.67-1.46) |
| 20+ | 287 | 80 | **0.59 (0.44-0.81)** | 73 | 0.83 (0.60-1.17) | **0.69 (0.53-0.89)** | 35 | 0.79 (0.50-1.24) |
| Don't know | 67 | 9 | **0.29 (0.14-0.59)** | 34 | **1.66 (1.04-2.66)** | 0.83 (0.55-1.26) | 7 | 0.67 (0.29-1.54) |
| Lab size |  |  |  |  |  |  |  |  |
| <10 | 417 | 182 | 1.00 | 171 | 1.00 | 1.00 | 80 | 1.00 |
| 10+ | 1222 | 388 | **0.73 (0.59-0.90)** | 425 | 0.85 (0.69-1.05) | **0.79 (0.66-0.93)** | 168 | **0.72 (0.54-0.96)** |
| Guidelines |  |  |  |  |  |  |  |  |
| N American | 718 | 271 | 1.00 | 276 | 1.00 | 1.00 | 102 | 1.00 |
| British | 337 | 84 | **0.66 (0.50-0.87)** | 129 | 1.00 (0.78-1.27) | 0.83 (0.68-1.02) | 38 | 0.79 (0.54-1.18) |
| Japanese | 98 | 56 | **1.51 (1.06-2.16)** | 11 | **0.29 (0.15-0.55)** | 0.90 (0.65-1.25) | 9 | 0.65 (0.32-1.32) |
| Other | 486 | 159 | 0.87 (0.69-1.09) | 180 | 0.96 (0.77-1.20) | 0.92 (0.77-1.09) | 99 | **1.43 (1.06-1.94)** |

**Supplementary table 8:**
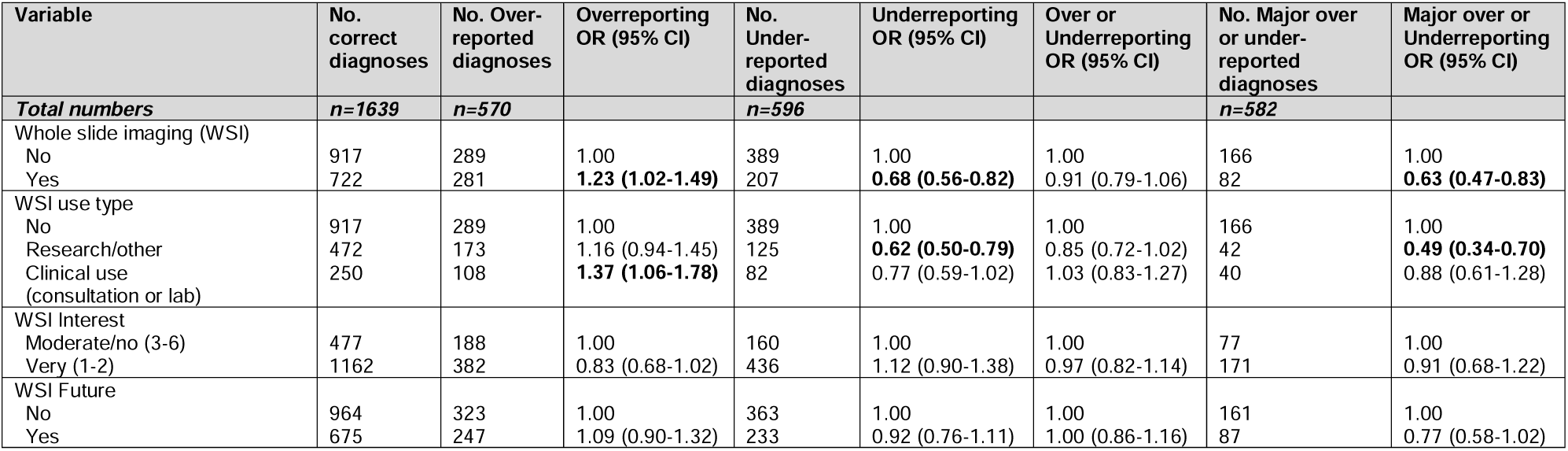
(c) Pathologist use and perceptions of whole slide imaging and odds of over or under-interpreting Barrett’s dysplasia: unadjusted analysis.

